# Association of viral variant and vaccination status with the occurrence of symptoms compatible with post-acute sequelae after primary SARS-CoV-2 infection

**DOI:** 10.1101/2022.10.21.22281349

**Authors:** Christian R. Kahlert, Carol Strahm, Sabine Güsewell, Alexia Cusini, Angela Brucher, Stephan Goppel, J. Carsten Möller, Manuela Ortner, Markus Ruetti, Reto Stocker, Danielle Vuichard-Gysin, Ulrike Besold, Allison McGeer, Lorenz Risch, Andrée Friedl, Matthias Schlegel, Pietro Vernazza, Stefan P. Kuster, Philipp Kohler, SURPRISE Study Group

**Affiliations:** Cantonal Hospital St Gallen, Division of Infectious Diseases and Hospital Epidemiology, St Gallen, Switzerland; Children’s Hospital of Eastern Switzerland, Department of Infectious Diseases and Hospital Epidemiology, St. Gallen, Switzerland; Cantonal Hospital of Grisons, Division of Infectious Diseases, Chur, Switzerland; Psychiatry Services of the Canton of St. Gallen (South), Switzerland; Psychiatry Services of the Canton of St. Gallen (North), Switzerland; Center for Neurological Rehabilitation, Zihlschlacht, Switzerland; Rheintal Werdenberg Sarganserland Hospital Group, Grabs, Switzerland; Fuerstenland Toggenburg Hospital Group, Wil, Switzerland; Hirslanden Clinic, Zurich, Switzerland; Thurgau Hospital Group, Division of Infectious Diseases and Hospital Epidemiology, Muensterlingen, Switzerland; Swiss National Centre for Infection Prevention (Swissnoso), Berne, Switzerland; Geriatric Clinic St. Gallen, St. Gallen, Switzerland; Sinai Health System, Toronto, Canada; Labormedizinisches Zentrum Dr Risch Ostschweiz AG, Buchs, Switzerland; Private Universität im Fürstentum Liechtenstein, Triesen, Liechtenstein; Center of Laboratory Medicine, University Institute of Clinical Chemistry, University of Bern, Inselspital, Bern, Switzerland; Cantonal Hospital Baden, Division of Infectious Diseases and Hospital Epidemiology, Baden, Switzerland

**Keywords:** SARS-CoV-2, health care workers, viral variants, omicron, post-acute, post-covid condition

## Abstract

**Importance:** Disentangling the effects of different SARS-CoV-2 variants and of vaccination on the occurrence of post-acute sequelae of SARS-CoV-2 (PASC) is crucial to estimate and potentially reduce the future burden of PASC.

**Objective:** To determine the association of primary SARS-CoV-2 infection on the frequency of PASC symptoms by viral variant and vaccination status.

**Design:** Cross-sectional questionnaire and SARS-CoV-2 serology (May/June 2022) performed within a prospective healthcare worker cohort (SURPRISE study).

**Setting:** Multicenter study in nine healthcare networks from North-Eastern Switzerland.

**Participants:** Volunteer sample of healthcare workers (HCW) from participating institutions. Of approximately 20’000 eligible participants, 3’870 registered for the cohort and 2’912 were included in this analysis.

**Exposures:** SARS-CoV-2 infection documented by positive nasopharyngeal swab (>4 weeks ago), stratified by viral variant and vaccination status at time of infection, compared to absence of documented infection (no positive swab, negative serology).

**Main Outcome:** Sum score of eighteen self-reported PASC symptoms.

**Results:** Among 2’912 participants (median age 44 years, 81.3% female), SARS-CoV-2 infection was reported by 1’685 (55.9%) participants, thereof 315 (18.7%) during Wild-type, 288 (17.1%) during Alpha/Delta, and 1’082 (64.2%) during Omicron circulation. Mean symptom number in previously infected participants significantly exceeded that of uninfected controls (0.39), but decreased with recency of the viral variant: 1.12 (p<0.001) for Wild-type (median time since infection 18.5 months), 0.67 (p<0.001) for Alpha/Delta (6.6 months), and 0.52 (p=0.005) for Omicron BA.1 (3.1 months) infected participants. After Omicron BA.1 infection, the mean symptom score was 0.49 (p=0.30) for those with ≥3 prior vaccinations and 0.71 (p=0.028) with 1-2 previous vaccinations compared to 0.36 for unvaccinated individuals. Adjusting for confounders, Wild-type (adjusted risk ratio [aRR] 2.81, 95% confidence interval [CI] 2.08-3.83) and Alpha/Delta infection (aRR 1.93, 95% CI 1.10-3.46) showed significant associations with the outcome, whereas Omicron BA.1 infection (aRR 1.29, 95% CI 0.69–2.43) and vaccination before infection (aRR 1.27, 95% CI 0.82–1.94) did not.

**Conclusions and Relevance:** Previous infection with pre-Omicron variants was the strongest risk factor for reporting PASC symptoms in this HCW cohort. A definite influence of prior vaccination on the prevention of PASC after Omicron BA.1 infection was not measurable.

## INTRODUCTION

Early in the current severe acute respiratory syndrome coronavirus-2 (SARS-CoV-2) pandemic, it has become apparent that not all patients fully recover from Coronavirus Disease 2019 (COVID-19), but have persistent physical and mental health symptoms^1,2^. As SARS-CoV-2 continues to evolve and its variants spread across the globe, post-acute sequelae after SARS-CoV-2 (PASC) or Long COVID represents a significant challenge to healthcare systems^3^. The estimates on the frequency of Long COVID vary greatly, depending on definitions used and time elapsed since infection^4,5^. The currently most common SARS-CoV-2 variant, Omicron, spreads even more efficiently but seems less pathogenic in acute disease, at least in a population with robust immunity^6^. As severity of acute infection has directly been associated with the risk of developing PASC^7,8^, this condition might be less common after Omicron infection compared to previous variants. Indeed, previous data show that compared to the Delta variant, after COVID-19 with Omicron, the risk for PASC is reduced by around 50%^9^. However, in this study, comparison with Alpha and the Wild-type was not possible; also, the study lacked a control group, which is crucial to correctly attribute these often non-specific symptoms to COVID-19^10^. Decreasing prevalence of PASC over time has also been reported from an Italian healthcare worker (HCW) cohort. In addition, the authors suggested that PASC develops less frequently in individuals who are vaccinated against SARS-CoV-2 before being infected^11^. Again, this study did not include a control group and infections occurring in the Delta and Omicron period were not evaluated separately.

In this study, we aimed to compare symptoms compatible with PASC between HCW after infection with different SARS-CoV-2 variants and uninfected seronegative controls. Also, we assessed the impact of mRNA COVID-19 vaccine on symptoms compatible with PASC.

## METHODS

### Setting and participants

The study was approved by the ethics committee of Eastern Switzerland. Written informed consent was obtained from participants. The observational multicenter study included volunteer HCW from eight healthcare networks in Northern/Eastern Switzerland^12^. The study population consisted mostly of participants from a previous HCW cohort, which had been launched in July/August 2020. In addition, new participants were recruited for the current study (**Figure S1**).

### Study procedures

Participants from the original cohort were prospectively followed between July/August 2020 and May/June 2022 with electronic questionnaires capturing SARS-CoV-2 positive swabs and vaccinations; also, repetitive SARS-CoV-2 serologies were performed (**Figure S2**)^13^. In May/June 2022, the cross-sectional study was performed including SARS-CoV-2 serology and an electronic questionnaire, asking about personal characteristics and risk factors, such as comorbidities, medications, household members, work type, exposure to COVID-19 patients, and the presence of PASC symptoms Participants also reported dates of previous SARS-CoV-2-positive nasopharyngeal swabs and vaccinations. Participants with multiple positive swabs and those with a first positive swab within 4 weeks or a vaccination within 1 week prior to the questionnaire were excluded.

### SARS-CoV-2 diagnostics

Participants were asked to get tested for SARS-CoV-2 in case of compatible symptoms, according to national recommendations. Depending on the participating institution, SARS-CoV-2 was detected by rapid antigen test or polymerase chain reaction. Self-reported nasopharyngeal swabs results were validated as previously described^13^. In May/June 2022, participants were serologically screened for anti-spike (anti-S) and anti-nucleocapsid (anti-N) antibodies, to identify pauci-/asymptomatically infected individuals^12^. The Roche Elecsys (Roche Diagnostics, Rotkreuz, Switzerland) electro-chemiluminescence immunoassay was used^14^.

### Definition of viral variant and vaccination status

The viral variant infecting a participant was inferred from the time of first positive swab according to sequencing data from Northeastern Switzerland^15^: Wild-type infections (February 2020 to January 2021); Alpha variant (February to June 2021); Delta variant (July to December 2021); Omicron variant (B.1.1.529.1; BA.1) (January to June 2022) (**Figure S2**). Infections during the Alpha period were merged with those during the Delta period due to the small number of Alpha infections. Participants without any previous positive SARS-CoV-2 swab and with negative anti-N in May/June 2022 were considered as uninfected controls. Those without positive SARS-CoV-2 swab but positive anti-N were excluded from further analyses. Vaccination status was assessed based on self-reported participant data. Over 99% of vaccinated participants indicated receipt of either the mRNA-1273 or the BNT162b2 vaccine.

### Outcomes

Participants were given a list of symptoms compatible with PASC and were asked to check those present at time of the questionnaire for more than 7 days but not before the pandemic. Symptoms included loss of smell/taste, shortness of breath, chest pain, hair loss, brain fog, tiredness/weakness, skin rash, muscle/limb pain, joint point, headache, nausea/anorexia, dizziness, stomachache, diarrhea, burnout/exhaustion, fever >38° Celsius or feverish feeling, chills, and cough. The main outcome was the total number of these 18 symptoms; individual symptoms were also analyzed.

As additional outcomes, participants answered a 9-item fatigue score (Fatigue Severity Scale, FSS)^16^, reported their self-rated health (SRH) on a 5-point scale (from “poor” to “excellent”)^17^, and indicated whether they think they would suffer from Long COVID (yes vs. no). For those answering the latter question with yes, the severity of Long COVID was assessed using the Post-COVID-19 Functional Status Scale (PCFS), a 5-point scale from “no restrictions in daily live” to “severe restrictions” (**Table S1**)^18,19^.

### Statistical analysis

We used descriptive statistics to compare baseline characteristics between those infected with the Wild-type virus, the Alpha/Delta variant, the Omicron BA.1 variant, and uninfected controls. For the main analysis, mean PASC symptom scores were compared between each of the infected groups and controls, respectively, using univariable negative-binomial models. Mean sum scores and 95% likelihood ratio confidence intervals (CI) were derived from models without intercept. The significance of differences was evaluated using Wald tests on coefficients of models with uninfected controls as intercept. The same analyses were performed separately by vaccination status: unvaccinated, vaccinated after infection (controls with any vaccination date), and vaccinated before infection with ≥1 vaccine dose (controls with any vaccination date). Second, the frequency of individual symptoms was compared between each of the infected groups and controls, respectively, using logistic regression. This analysis was performed separately for those unvaccinated at time of infection (ie, never vaccinated individuals or vaccination after infection) and those with ≥1 vaccine dose before infection. Controls were uninfected individuals without and with any vaccination, respectively.

Third, we compared the mean symptom score between boosted (≥3 vaccinations at least 7 days before infection) and non-boosted (1-2 vaccinations) compared to unvaccinated (but also infected) participants using negative-binomial models. This analysis was restricted to participants infected in the Delta (without the Alpha group) and Omicron period, because booster vaccination was not available before.

### Multivariable analysis

To assess the independent impact of viral variant and vaccination status on symptom number, we used a multivariable negative-binomial regression model. Variables previously found to be associated with the occurrence of Long COVID were included in the model (**Table S1**)^7,20^. Rate ratios (RR) derived from model coefficients and corresponding 95% CI are shown as forest plots; P-values were obtained from Wald tests.

### Sensitivity analysis

To minimize the risk of incorrect group assignment due to undetected earlier SARS-CoV-2 infection, we re-ran the main analysis and the multivariable analysis including only individuals with previous serology data available and excluding those with any positive anti-N before the first positive SARS-CoV-2 swab or before the survey (for controls) (**Figure S1**).

### Analysis of additional outcomes

Additional outcomes were analyzed between each of the infected group and controls. For this purpose, we used a negative-binomial model (FSS), logistic regression (self-classification of having Long COVID), and proportional-odds logistic regression (distribution of SRH ratings and PCFS). These analyses were not stratified by vaccination status. R statistical software Version 4.0.2 was used for statistical analysis; two-sided p-values of <.05 were considered significant. This report follows the STROBE reporting guideline for observational studies^21^.

## RESULTS

### Study population and SARS-CoV-2 infections

Of approximately 19’600 eligible study participants, 3’870 answered the questionnaire in May/June 2022 and 2’912 were included in the analysis (**Figure S1**). Median age was 44 years and 81.3% were women. SARS-CoV-2 infection was reported by 1’685 (57.9%) participants, thereof 315 (18.7%) during the Wild-type, 288 (17.1%) during the Alpha/Delta, and 1’082 (64.2%) during the Omicron period. Median time since infection was 18.3 months (interquartile range [IQR] 17.5-19.2) for Wild-type, 6.5 (IQR 6.0-9-0) for Alpha/Delta, and 3.1 (IQR 2.6-4.0) for Omicron BA.1 infections. The group of uninfected controls consisted of 1’227 individuals. Of the 2’912 individuals, 2’570 reported baseline characteristics (**Table 1)**.

**Table 1.** Characteristics of 2’570 participants answering the study questionnaire by time of first positive SARS-CoV-2 swab.

| Baseline characteristics | Wild-type infection<br>*n* = 283 | Alpha/Delta infection<br>*n* = 268 | Omicron infection<br>*n* = 963 | No infection<br>*n* = 1056 | *p*-value<sup>b</sup> |
| --- | --- | --- | --- | --- | --- |
| Age (years, median and IQR) | 44 (33-54) | 42 (32-49) | 42 (34-52) | 48 (37-56) | <0.001 |
| Sex (male vs. female) | 54 (19.1%) | 47 (17.5%) | 173 (18.0%) | 206 (19.5%) | 0.787 |
| BMI (>30 vs. ≤30 kg/m<sup>2</sup>) | 40 (14.1%) | 23 (8.6%) | 97 (10.1%) | 134 (12.7%) | 0.055 |
| Ethnicity (Caucasian) | 275 (97.2%) | 259 (96.6%) | 946 (98.2%) | 1024 (97.0%) | 0.442 |
| Child ≤ 6 years in household | 36 (12.7%) | 36 (13.4%) | 150 (15.6%) | 91 (8.6%) | <0.001 |
| Any comorbidity | 143 (50.5%) | 116 (43.3%) | 467 (48.5%) | 514 (48.7%) | 0.343 |
| Pollen allergy | 86 (30.4%) | 77 (28.7%) | 322 (33.4%) | 331 (31.3%) | 0.438 |
| Other comorbidities | 86 (30.4%) | 58 (21.6%) | 254 (26.4%) | 299 (28.3%) | 0.085 |
| Any medication | 97 (34.3%) | 66 (24.6%) | 271 (28.1%) | 335 (31.7%) | 0.026 |
| Active smoking | 41 (14.5%) | 44 (16.4%) | 154 (16.0%) | 200 (18.9%) | 0.186 |
| Alcohol (>1 drink per week) | 113 (39.9%) | 103 (38.4%) | 424 (44.0%) | 419 (39.7%) | 0.089 |
| Profession |  |  |  |  | <0.001 |
| Nurse | 175 (61.8%) | 149 (55.6%) | 459 (47.7%) | 476 (45.1%) |  |
| Physician | 27 (9.5%) | 23 (8.6%) | 127 (13.2%) | 163 (15.4%) |  |
| Other | 81 (28.6%) | 96 (35.8%) | 377 (39.1%) | 417 (39.5%) |  |
| Cumulative contact duration to COVID-19 patients (minutes, median and IQR) | 400 (30-2250) | 285 (1-1200) | 150 (1-1000) | 130 (1-750) | <0.001 |
| Intensive care | 52 (18.4%) | 32 (11.9%) | 133 (13.8%) | 140 (13.3%) | 0.113 |
| Full-time work (>0.8 FTE) | 147 (51.9%) | 128 (47.8%) | 475 (49.3%) | 561 (53.1%) | 0.239 |
| Vaccination<sup>a</sup> |  |  |  |  | <0.001 |
| before infection | 0 (0.0%) | 165 (61.6%) | 876 (91.0%) | 0 (0.0%) |  |
| after or without infection | 255 (90.1%) | 35 (13.1%) | 3 (0.3%) | 1022 (96.8%) |  |
| no vaccination | 28 (9.9%) | 68 (25.4%) | 84 (8.7%) | 34 (3.2%) |  |

IQR, Interquartile Range; BMI, Body Mass Index; FTE, Full Time Equivalent
<sup>a</sup> Note that numbers are slightly lower than in Figure 1 because 342 individuals only reported data on infections and vaccinations, but no further data.
<sup>b</sup> Global test for the significance of group differences. Kruskal-Wallis test for numeric variables (age, cumulative patient contact duration) and Chi-squared test for categorical variables (all others).

### PASC symptoms and viral variant

The 2’912 participants reported a mean of 0.55 symptoms (including those without symptoms). Mean symptom number in the three groups of previously infected participants significantly exceeded that of uninfected controls (0.39), but decreased with recency of the viral variant: 1.12 (p<0.001) for Wild-type, 0.67 (p<0.001) for Alpha/Delta, and 0.52 (p=0.005) for Omicron BA.1 infected participants (**Figure 1a**). Similar decreasing trends across viral variants were observed for unvaccinated participants (**Figure 1b**), for those with vaccination before or after infection (**Figure 1c**), and also in the sensitivity analysis, although sample size was small in individual subgroups (**Figure S3**).

**Figure 1.**
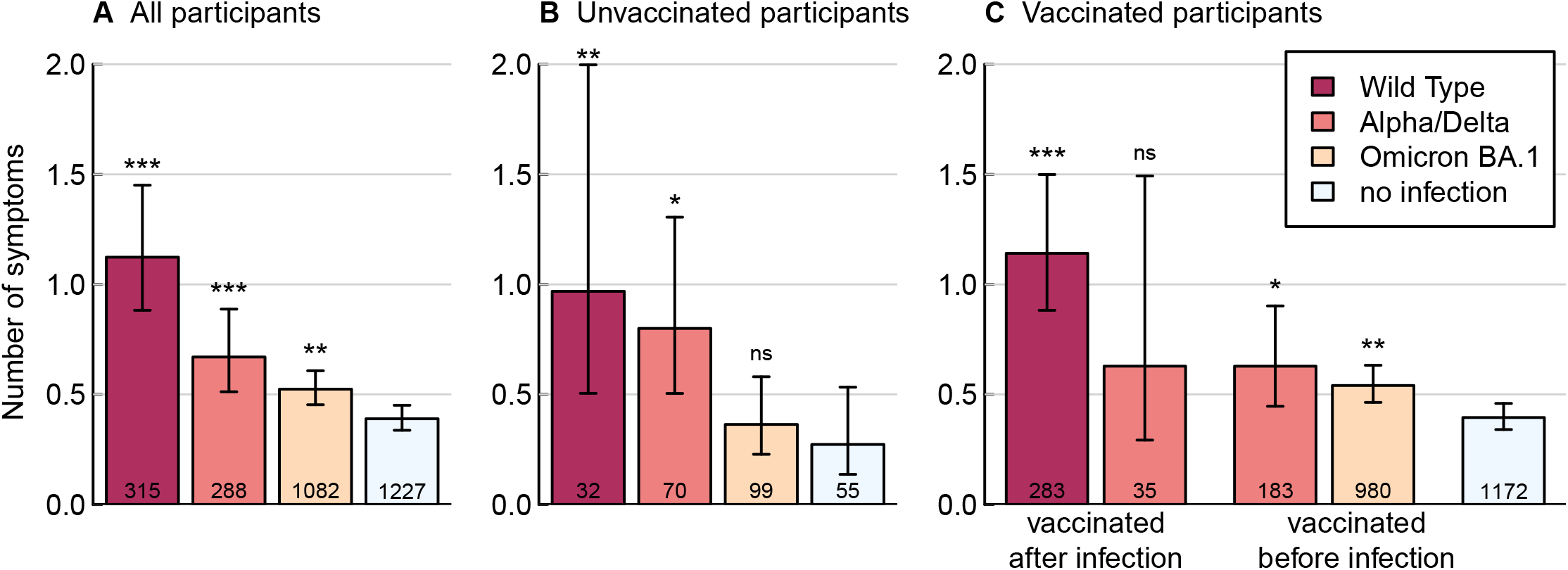
Cross-sectional analysis of May/June 2022. Means and 95% confidence intervals of post-acute sequelae of SARS-CoV-2 symptom score by viral variant and vaccination status. Asterisks above bars indicate statistical significance in reference to uninfected participants with same vaccination status, respectively, as obtained through Wald tests on coefficients of negative binomial models with uninfected participants as reference group (***, p < 0.001; **, p < 0.01; *, p < 0.05; ^ns^, p ≥ 0.05). N at the bottom of the bars designate number of participants. Note: there were no vaccinations before Wild-type infection and only three individuals vaccinated after Omicron BA.1 infection (not shown in panel C).

At least one symptom was reported by 695 (23.9%) participants, and this share was significantly raised after infection: 38.7% for Wild-type infected participants (p<0.001), 31.6% for Alpha/Delta (p=0.001), 23.8% for Omicron BA.1 (p=0.001), compared to 18.3% for uninfected individuals. The most commonly reported symptoms overall were tiredness/weakness with a frequency of 14.7% in infected and 9.4% in uninfected participants. Symptoms consistently associated with SARS-CoV-2 infection across all variants were loss of smell/taste and hair loss for unvaccinated (**Figure S4**), and loss of smell/taste and brain fog for vaccinated participants (**Figure S5**).

### PASC symptoms and vaccination status

Among participants infected during the Delta period, vaccinated individuals (+/-booster) reported on average fewer symptoms than unvaccinated participants; however, the differences were not statistically significant. After Omicron BA.1 infection, the mean reported symptom number was 0.49 (p=0.30) for those with ≥3 prior vaccinations and 0.71 (p=0.028) for those with 1-2 previous vaccinations compared to 0.36 for unvaccinated individuals (**Figure 2**).

**Figure 2.**
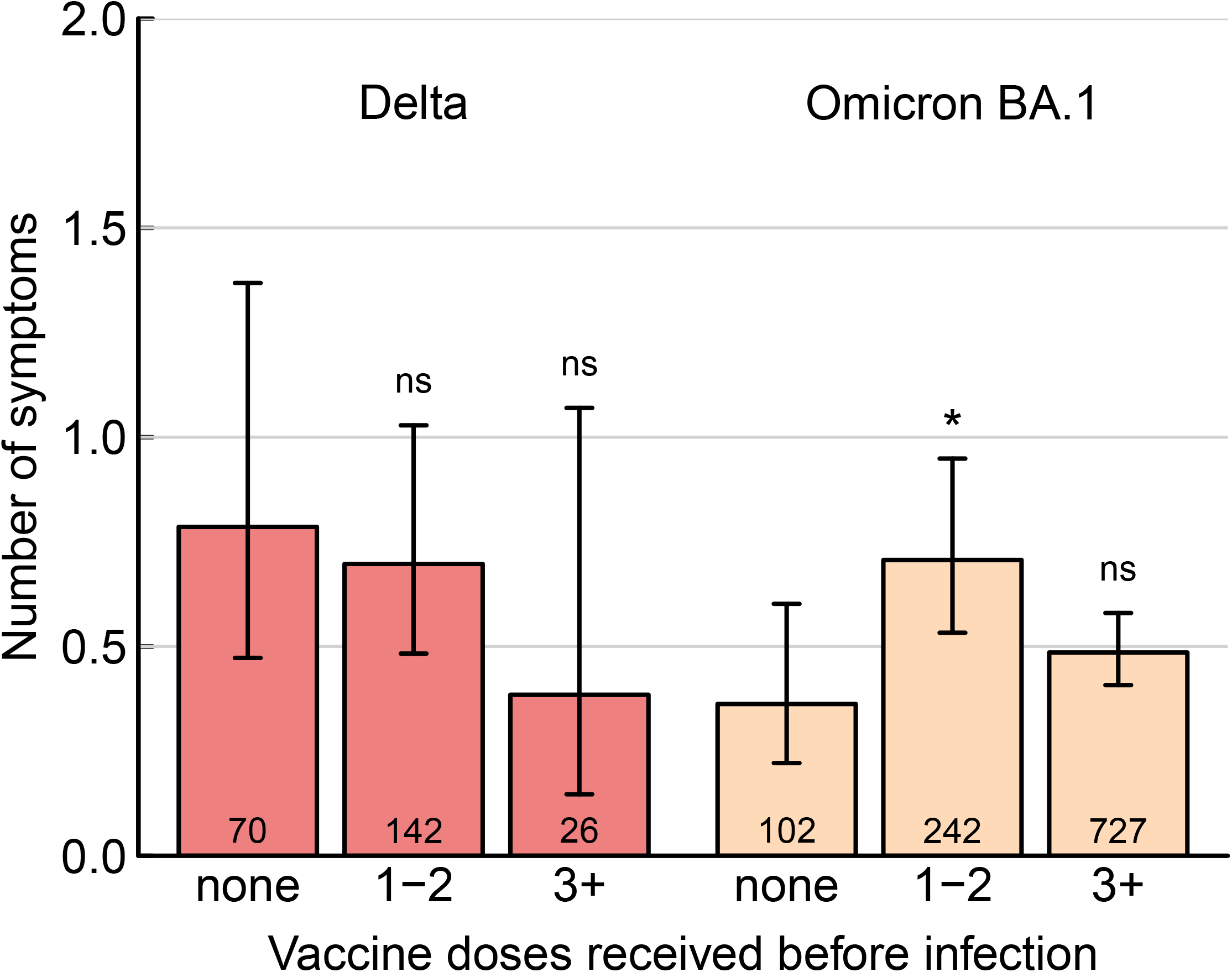
Means and 95% confidence intervals of post-acute sequelae of SARS-CoV-2 symptom score in relation to number of vaccine doses received before positive swab. Left: Participants after infection in Delta period (1 July 1^st^ to December 31^th^ 2021). Right: Participants after infection in Omicron BA.1 period (January 1^st^ to June 30^th^ 2022). Asterisks above bars indicate statistical significance in reference to unvaccinated participants infected in the same period, respectively, as obtained through Wald tests on coefficients of negative binomial models with group “none” as reference (*, p < 0.05; ^ns^, p ≥ 0.05).

### Multivariable analysis

In multivariable analysis, infection with the Wild-type virus (adjusted rate ratio [aRR] 2.81, 95% CI 2.08–3.83) and with the Alpha/Delta variant (aRR 1.93, 95% CI 1.10–3.46) were positively associated with the number of symptoms reported, whereas infection during the Omicron period (aRR 1.29, 95% CI 0.69–2.43) and vaccination before infection (aRR 1.27, 95% CI 0.82–1.94) were not. Other variables associated with the outcome were body mass index (BMI) >30m/kg^2^ (aRR 1.43, 95% CI 1.08–1.92), having a pre-existing comorbidity (RR 1.35, 95% CI 1.11–1.65), being on any medication (aRR 1.49, 95% CI 1.20–1.86) and cumulative COVID-19 patient contact (aRR 1.11, 95% CI 1.01–1.21) (**Figure 3, Table S2**). Effect sizes for viral variants increased slightly in sensitivity analysis, whereas other co-variables, except being on any medication, were no longer associated with the outcome (**Figure S6**).

**Figure 3.**
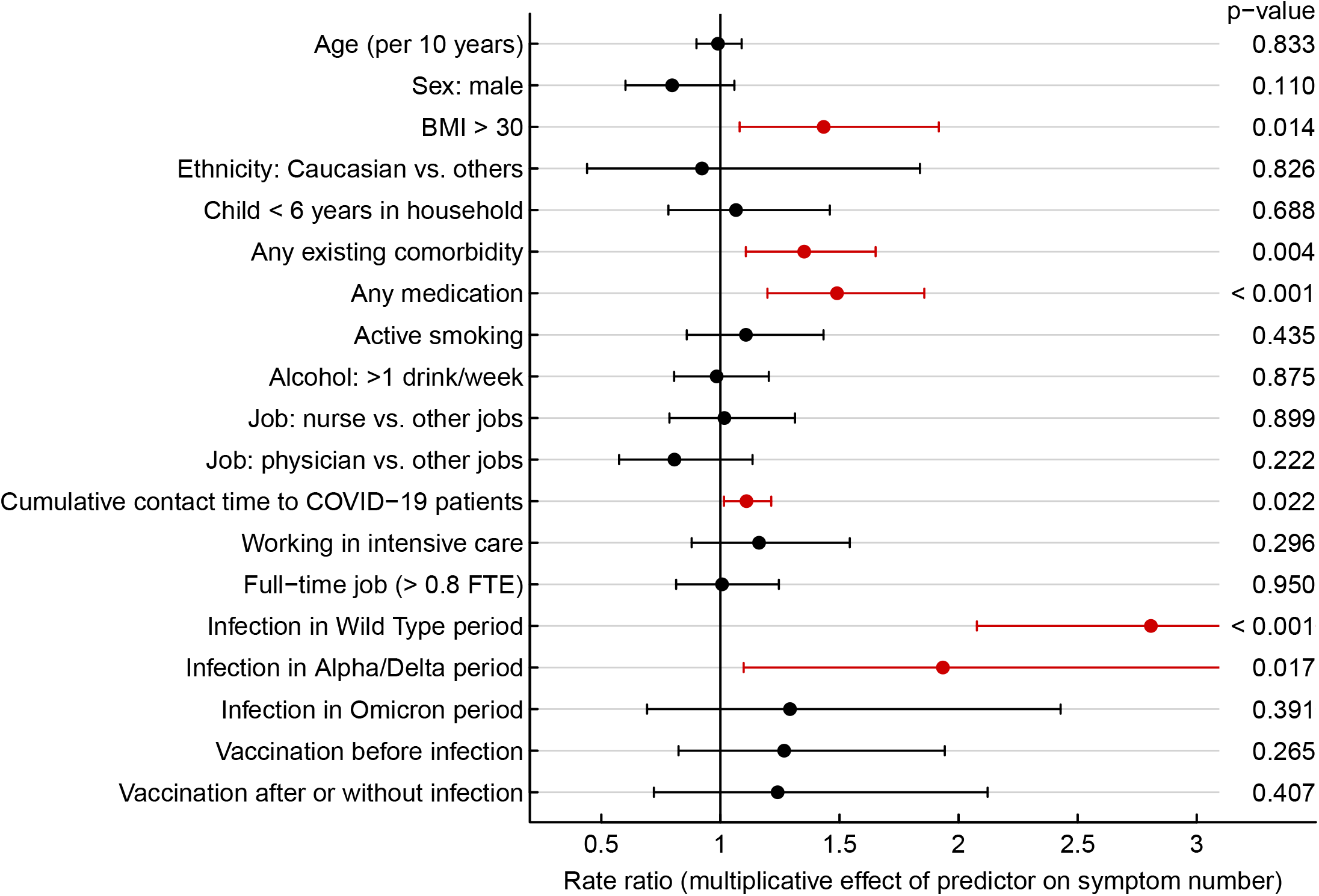
Results of multivariable negative binomial model regarding number of symptoms compatible with post-acute sequelae of SARS-CoV-2. Predictors positively associated with symptom number (i.e. 95% confidence interval for rate ratio entirely above 1) are highlighted in red. For numeric values of point estimates and 95% confidence intervals see Table S3.

### Additional outcomes

Both the FSS and the SRH were highest or lowest, respectively, for those after Wild-type infection, while no difference was observed between those with Alpha/Delta or Omicron BA.1 infection and controls (**Table 2**). The percentage of participants who considered themselves suffering from Long COVID was substantially higher after Wild-type (17.1%, p<0.001), Alpha/Delta (10.4%, p<0.001), and Omicron BA.1 infection (4.8%, p<0.001) compared to controls (0.9%). For those suffering from Long COVID, functional impairment in daily live as per the PCFS score was similar between groups.

**Table 2.**
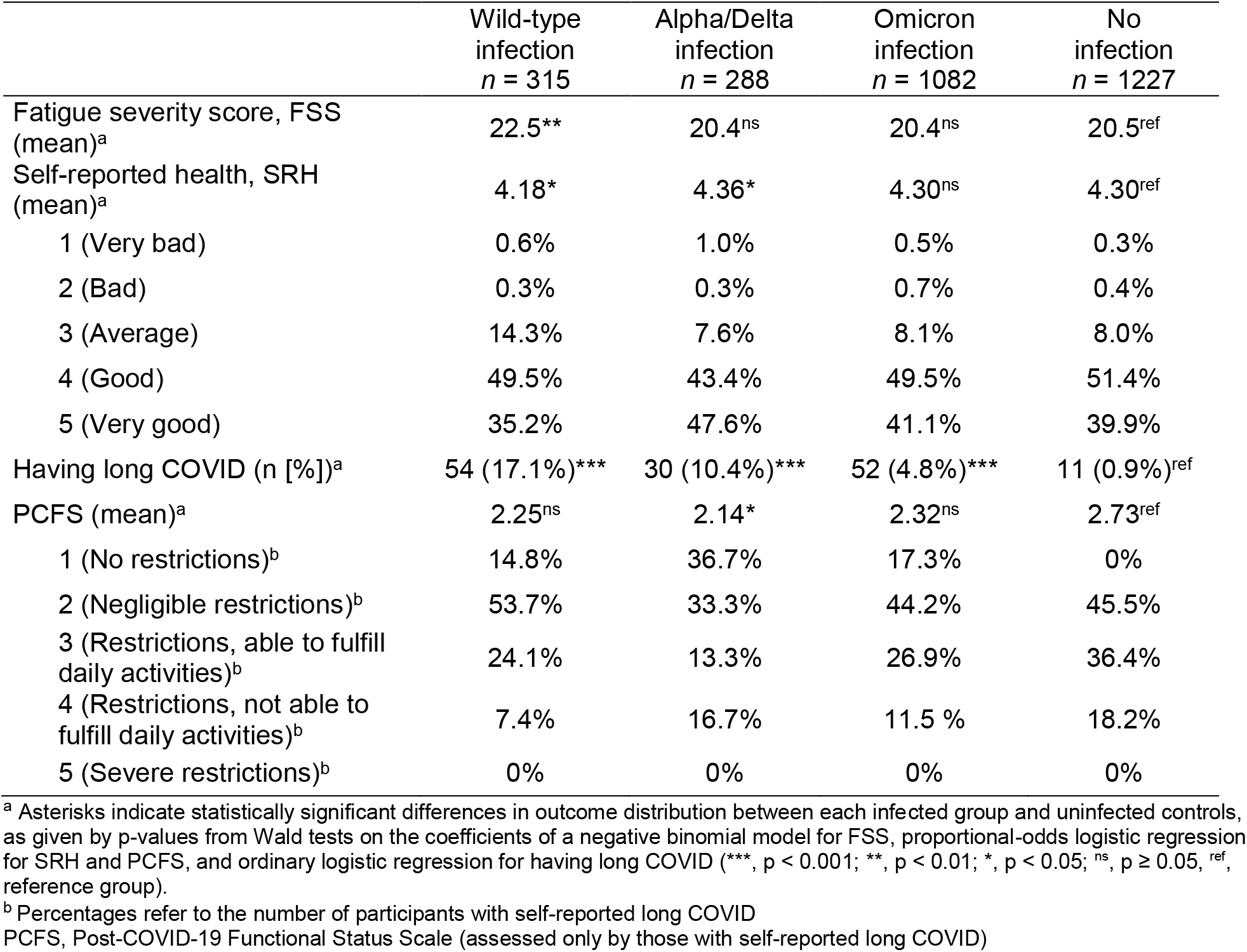
Results of additional outcomes by time of first infection (or no infection) among 2’912 participants.

## DISCUSSION

Within a well-defined HCW population, we show that the most decisive risk factor for reporting symptoms compatible with PASC is the viral variant causing the primary infection. Participants after Wild-type SARS-CoV-2 and Alpha/Delta infections reported the highest numbers even 18 and 6 months after infection, respectively. In contrast, those at 3 months after Omicron BA.1 infection are only minimally affected compared to uninfected controls. SARS-CoV-2 vaccination including receipt of booster vaccine before Omicron BA.1 infection was not associated with less PASC symptoms.

The viral variant was the most important risk factor associated with the presence of PASC symptoms, even after adjusting for co-variables typically associated with Long COVID, such as age, sex, BMI, and vaccination status. Individuals after Wild-type reported significantly more PASC symptoms, had a higher fatigue score and a lower SRH than uninfected controls. These findings are particularly worrying considering that Wild-type infections occurred a median of 18 months before this survey. Few studies have reported such a long duration of PASC. Persistence of symptoms for 12 months have been reported by several others, both in hospitalized and non-hospitalized populations^22–24^. One study demonstrated the persistence of memory problems at 18 months after mild SARS-CoV-2 infection and another showed that 38% of infected patients still reported symptoms at 23 months after infection^25,26^. In addition, some authors have suggested that loss of smell/taste, which was one of the symptoms showing a clear association with PASC in our study, might potentially be a permanent sequela after COVID-19^27^. Further research aiming at mitigating the PASC burden in those infected early on in the pandemic is urgently needed.

The frequency of Long COVID after infection with the Omicron variant has been reported to be 5% at ≥4 weeks after infection, which is about half compared to the Delta variant^9^. Comparing people infected with Omicron vs. previous variants, 5 vs. 55% reported ≥1 PASC symptom in another study^28^. However, both studies have not included a non-infected control group, which allows to better interpret the significance of these findings. In our study, participants after Omicron BA.1 infection still reported more symptoms than uninfected controls. However, we found that many symptoms typically associated with PASC, such as fatigue, exhaustion or muscle/limb pain,^1^ were only slightly or not at all more common after Omicron BA.1 infection compared to uninfected controls. On the other hand, the high prevalence of some of these symptoms in uninfected individuals highlights the importance to also consider other causes, besides COVID-19, underlying these symptoms. This particularly includes conditions such as chronic fatigue syndrome/myalgic encephalomyelitis, which have been shown to be often related to other viral infections^29–31^. Importantly, in contrast to participants after Wild-type infection, FSS and SRH were not any different between Omicron BA.1 infected and uninfected controls.

Our data further suggest that vaccination with SARS-CoV-2 mRNA vaccines prior to Omicron BA.1 infection does not lower the risk of PASC. This is in contrast to previous studies from the pre-Omicron era, which showed a benefit of vaccination against PASC^32,33^. In addition, an Italian study found that vaccination before infection during the Delta/Omicron period was indeed associated with decreased risk for long COVID^11^. The discrepancy can be explained by the fact that in this study, infections during the Delta and Omicron period were pooled; hence, the effect of previous vaccination on Long COVID could have been mainly driven by Delta infections. In line with this hypothesis, our data suggest that previous vaccination might have been protective against PASC after Delta, although results were not statistically significant due to the small number of cases. In contrast, previous vaccination was clearly not associated with a reduction of PASC symptoms after Omicron BA.1 infection despite the large number of cases. Further studies are needed to confirm our finding.

Important strengths of our study are the clearly defined infection and vaccine status of every individual study participant as well as the relatively large share of hitherto unvaccinated participants. This enabled us to adequately disentangle the effects of viral variant and of SARS-CoV-2 vaccination on the occurrence of PASC symptoms, a task which is increasingly becoming unrealizable due to high vaccination and infection rates, particularly in HCW^34^. The probably most important strength is the inclusion of a non-infected control group, which many previous studies on Long COVID have lacked^10^. A Dutch study recently reported the frequency of PASC symptoms in infected vs. non-infected individuals, and attributed these symptoms in 12.7% of cases to COVID-19. Yet, the study was performed before circulation of the Delta and Omicron variants; furthermore, no SARS-CoV-2 serology was performed which would allow to also capture asymptomatic infections^35^. We relied on anti-N negativity to exclude previous infection. We acknowledge that this definition may have missed a certain proportion of infected individuals due to lack of seroconversion or waning antibody titers over time^14^. In a post-hoc analysis, we found that 11.7% of those with negative anti-N reported indeed a positive swab. By considering both criteria, anti-N negativity and lack of positive swab, we think that our approach of defining an uninfected control group is reasonable. Furthermore, results of the sensitivity analysis, which also considered previous serology results and excluded those with waning immunity over time, largely confirmed our main findings.

Our study has further limitations. First, definition of viral variants was based on population data and not on individual sequencing data. This might have led to misclassifications of viral variants in any direction and therefore to an underestimation of the observed differences. Second, the choice of our outcome definition can be debated. The large number of symptoms and the less strict time criterion (symptoms had to be present at least for 7 days) compared to other studies might have led to an overestimation of PASC prevalence. Third, we cannot exclude that vaccine hesitancy could be associated with the way people perceive and/or report physical and mental health symptoms, which would distort the comparability between vaccinated and unvaccinated participants. Fourth, our data lack generalizability both in terms of study population as well as of viral variants. We cannot exclude that Omicron BA.1 infections occurring in the elderly or more comorbid populations, where acute infection might manifest itself more severely, might confer an increased risk of PASC; also, our results are not necessarily applicable to newer Omicron variants.

To conclude, these data suggest that in a HCW population of predominantly young, healthy Caucasian females, individuals infected with the Wild-type, and less so with the Alpha/Delta variants, still report symptoms compatible with PASC after a median of 18 and 6 months, respectively, whereas the burden at 3 months after Omicron BA.1 infection is considerably lower. Vaccination prior to Omicron BA.1 infection was not associated with less PASC symptoms. Future research should primarily address the needs of individuals infected early on in the pandemic, whereas in individuals suffering from symptoms compatible with PASC after Omicron BA.1 infection, other aetiologies besides COVID-19 should be actively sought.

## Data Availability

All data produced in the present study are available upon reasonable request to the authors

## Acknowledgments

We also thank the participants of the SURPRISE study and the members of the study group (in alphabetical order): Ulrike Besold, MD (Geriatric Clinic St. Gallen), Angela Brucher, MD (Psychiatry Services South, St. Gallen), Alexia Cusini, MD (Cantonal Hospital Graubünden), Thomas Egger (Cantonal Hospital St. Gallen), Andrée Friedl, MD (Cantonal Hospital Baden), Stephan Goppel, MD (Psychiatry Services North, St. Gallen), Fabian Grässli, MSc (Cantonal Hospital St. Gallen), Christian R. Kahlert, MD (Children’s Hospital of Eastern Switzerland, St. Gallen), Joelle Keller (Hirslanden Clinic Zurich), Simone Kessler (Cantonal Hospital St. Gallen), Philipp Kohler, MD MSc (Cantonal Hospital St. Gallen), Stefan P. Kuster, MD MSc (Cantonal Hospital St. Gallen), Onicio Leal, PhD (University of Zurich), Eva Lemmenmeier, MD (Clienia Littenheid), Allison McGeer, MD MSc (Mount Sinai Hospital, Toronto), Dorette Meier Kleeb, MD (Cantonal Hospital Baden), J. Carsten Möller, MD (Clinic Zihlschlacht), Maja F. Müller (Hirslanden Clinic Zurich), Vaxhid Musa (Cantonal Hospital St. Gallen), Manuela Ortner (Rheintal Werdenberg Sarganserland Hospital Group, Grabs), Philip Rieder, PhD (Hirslanden Clinic Zurich), Lorenz Risch, MD PhD (Laboratory Risch Buchs), Markus Ruetti, MD (Fuerstenland Toggenburg Hospital Group Wil), Matthias Schlegel, MD (Cantonal Hospital St. Gallen), Hans-Ruedi Schmid, PhD (Cantonal Hospital Baden), Reto Stocker, MD (Hirslanden Clinic Zurich), Pietro Vernazza, MD (Cantonal Hospital St. Gallen), Matthias von Kietzell MD (Clinic Stephanshorn St. Gallen), Danielle Vuichard-Gysin, MD MSc (Thurgau Hospital Group Muensterlingen), and Benedikt Wiggli, MD (Cantonal Hospital Baden).

This work was supported by the Swiss National Sciences Foundation (grant number 31CA30_196544; grant number PZ00P3_179919 to PK), the Federal Office of Public Health (grant number 20.008218/421-28/1), the Health Department of the Canton of St. Gallen, and the research fund of the Cantonal Hospital of St. Gallen. Funding institutions did not have any role in design and conduct of the study; collection, management, analysis, and interpretation of the data; preparation, review, or approval of the manuscript; and decision to submit the manuscript for publication.

The authors have no other conflicts of interest to declare.

## Notes

### Competing Interest Statement

The authors have no conflicts of interest to declare.

### Author Declarations

Ethcis committee of Eastern Switzerland (EKOS) gave ethical approval for this work.

