## Supplementary material for "Association of viral variant and vaccination status with the occurrence of symptoms compatible with post-acute sequelae after primary SARS-CoV-2 infection"

**Kahlert et al.**

**SUPPLEMENTS**

**SUPPLEMENTARY TABLES**

Table S1. Co-variable definitions, levels, and time points when variables were obtained. All variables collected (for new participants) or updated (for former participants) in May 2022.

| **Variable name (unit)** | **Definition** | **Levels** |
| --- | --- | --- |
| Anthropometrics/baseline health |  |  |
| Age (years) | Age reached in 2022 | Number |
| Body mass index (kg/m2) | Body mass index at time of study entry | ≤30; >30 |
| Sex |  | Female; Male (Ref^a^) |
| Ethnicity | Self-reported ethnicity | Caucasian vs. others |
| Comorbidities | Presence of any of the following: arterial hypertension, diabetes, cancer, pulmonary disease, immune deficiency, hypertension, CVD, allergy, other | Yes; No (Ref) |
| Any medication | Taking any medication on a regular basis, except for contraception and vitamins or other preventive medication against infections. | Yes; No (Ref) |
| Active smoking | Self-reported active smoker | Yes; No (Ref) |
| Alcohol | Self-reported alcohol consumption in drinks/week | ≤1 drink (Ref); >1 drink |
| SARS-CoV-2 exposures and risk behaviors |  |  |
| Profession (job) | Current profession | Physician; Nurse; Other |
| Intensive care | Works partly or mostly on intensive or intermediate care unit | Yes; No (Ref) |
| Cumulative duration of COVID-19 patient contact | Number of COVID-19 patients seen in last 12 months; mean duration of these contacts. | log10 (patient number*mean contact time per patient in minutes) |
| Full-time equivalent (FTE) | Full-time equivalent in percent | ≤0.8 FTE (Ref); >0.8 FTE |
| SARS-CoV-2 vaccinations and infections |  |  |
| SARS-CoV-2 vaccination | First SARS-CoV-2 vaccination | Yes, before date of first SARS-CoV-2 infection; Yes, after date of infection or no infection; No (Ref) |
| SARS-CoV-2 infections | First positive SARS-CoV-2 swab reported during follow-up | According to date of positive swab: Wild-type infections (February 2020 to January 2021); Alpha variant (February 2021 to June 2021); Delta variant (July 2021 to December 2021); Omicron variant (B.1.1.529.1; BA.1) (January 2022 to June 2022) |
| Self-reported health |  |  |
| Health status | How would you rate your current health status? | 1, Very bad; 2, Bad; 3, Average; 4, Good; 5, Very good |
| Long COVID | Do you think that you currently suffer from long COVID (irrespective of previous documented infection)? | Yes; No (Ref) |
| Severity of long COVID | How much are you restricted in your daily living due to long COVID symptoms? | 1, No restrictions or symptoms, pain, depression or anxiety;  2, I do have negligible restrictions in my daily living, because I can participate at all usual daily activities despite persistent symptoms, pain, depression or anxiety;  3, I am suffering from restrictions in my daily living because I have to avoid or reduce certain acitivites due to symptoms, pain, depression or anxiety. I am capable of fulfilling my activities without support;  4, I am suffering from restrictions because I am not able to fulfill all my activity in my daily living due to symptoms, pain, depression or anxiety. I am capable of caring for myself without support;  5, I am suffering from severe restrictions in my daily living. I am not capable of caring for myself and I am dependent on others due to symptoms, pain, depression or anxiety. |

Ref, Reference

Table S2. Results of univariable and multivariable negative binomial model regarding number of reported post-Covid symptoms. RR, Rate Ratios; aRR, Adjusted Rate Ratio; CI, 95% Confidence Intervals, N = 2570 (except for missing values) in univariable analysis, N = 2452 in multivariable analysis.

|  | **Univariable analysis** | | | **Multivariable analysis** | |
| --- | --- | --- | --- | --- | --- |
|  | **RR, 95% CI** | **p-value** | **n missing** | **aRR, 95% CI** | **p-value** |
| Age (per decade) | 1.00 (0.92–1.09) | 0.998 | 0 | 0.99 (0.90–1.09) | 0.833 |
| Sex (male vs. female) | 0.73 (0.57–0.94) | 0.015 | 0 | 0.80 (0.60–1.06) | 0.110 |
| Body mass index (>30 vs. ≤30 kg/m^2^) | 1.67 (1.26–2.23) | <0.001 | 3 | 1.43 (1.08–1.92) | 0.014 |
| Ethnicity (Caucasian vs. others) | 0.85 (0.41–1.65) | 0.654 | 17 | 0.92 (0.44–1.84) | 0.826 |
| Any child ≤6 years in household | 0.98 (0.73–1.32) | 0.891 | 0 | 1.07 (0.78–1.46) | 0.688 |
| Any comorbidity | 1.56 (1.29–1.89) | <0.001 | 0 | 1.35 (1.11–1.65) | 0.004 |
| Any medication | 1.73 (1.41–2.12) | <0.001 | 0 | 1.49 (1.20–1.86) | <0.001 |
| Active smoking | 1.08 (0.84–1.39) | 0.568 | 0 | 1.11 (0.86–1.43) | 0.435 |
| Alcohol (>1 drink per week) | 0.83 (0.68–1.02) | 0.072 | 104 | 0.98 (0.81–1.20) | 0.875 |
| Profession |  |  |  |  |  |
| Nurse (vs. others) | 1.36 (1.11–1.67) | 0.004 | 0 | 1.02 (0.79–1.31) | 0.899 |
| Physician (vs. others) | 0.82 (0.59–1.13) | 0.214 | 0 | 0.81 (0.57–1.14) | 0.222 |
| Cumulative contact time (minutes) to Covid-19 patients (per 10-fold increase) | 1.16 (1.09–1.25) | <0.001 | 0 | 1.11 (1.01–1.21) | 0.022 |
| Work in intensive care | 1.44 (1.11–1.89) | 0.007 | 0 | 1.16 (0.88–1.54) | 0.296 |
| Full-time work (>0.8 vs. ≤0.8 FTE) | 0.95 (0.79–1.16) | 0.638 | 0 | 1.01 (0.81–1.25) | 0.950 |
| SARS-CoV-2 infection |  |  |  |  |  |
| Wild-type infection (vs. none) | 3.17 (2.36–4.29) | <0.001 | 0 | 2.81 (2.08–3.83) | <0.001 |
| Alpha/Delta infection (vs. none) | 1.81 (1.32–2.50) | <0.001 | 0 | 1.93 (1.10–3.46) | 0.017 |
| Omicron infection (vs. none) | 1.35 (1.09–1.68) | 0.007 | 0 | 1.29 (0.69–2.43) | 0.391 |
| Vaccination |  |  |  |  |  |
| Before infection (vs. no vaccination) | 1.04 (0.71–1.49) | 0.839 | 0 | 1.27 (0.82–1.94) | 0.265 |
| After/no infection (vs. no vaccination) | 1.05 (0.73–1.50) | 0.793 | 0 | 1.24 (0.72–2.12) | 0.407 |

FTE, Full Time Equivalent

**FIGURES**

Figure S1. Study flow of participants. Participants had either been included in the original prospective cohort or were newly recruited for this cross-sectional analysis.

Figure S2. Study procedures and timeline (prospective cohort and cross-sectional study) in relation to SARS-CoV-2 vaccine availability and number of reported COVID-19 cases in Switzerland per 100’000 population^1^. Predominating viral variant based on national sequencing data^2^.

Figure S3. Sensitivity analysis excluding individuals with positive anti-nucleocapsid antibodies before first reported infection and those without previous serology results. Cross-sectional analysis of May/June 2022. Means and 95% confidence intervals of post-Covid symptom score by time of positive swab. Asterisks above bars indicate statistical significance in reference to uninfected participants with same vaccination status, respectively (***, p < 0.001; **, p < 0.01; *, p < 0.05; no symbol, p ≥ 0.05).

Figure S4. Percentage and 95% confidence intervals of reported post-Covid symptoms by time of positive swab; all participants were unvaccinated at time of infection (but partly vaccinated after infection). Asterisks above bars indicate statistical significance in reference to uninfected participants (***, p < 0.001; **, p < 0.01; *, p < 0.05; no symbol, p ≥ 0.05).

Figure S5. Percentage and 95% confidence intervals of reported post-Covid symptoms by time of positive swab; all participants had received at least one vaccine dose at time of infection. Asterisks above bars indicate statistical significance in reference to uninfected participants (***, p < 0.001; **, p < 0.01; *, p < 0.05; no symbol, p ≥ 0.05). Note: Vaccine not available before Wild-Type infection.

Figure S6. Sensitivity analysis excluding individuals with positive anti-nucleocapsid antibodies before first reported infection and those without previous serology results. Results of multivariable negative binomial model regarding number of reported post-Covid symptoms. Factors positively associated with symptom number are highlighted in red.

**REFERENCES**

1.	*BAG Coronavirus, Situation Schweiz*. https://www.bag.admin.ch/bag/de/home/krankheiten/ausbrueche-epidemien-pandemien/aktuelle-ausbrueche-epidemien/novel-cov/situation-schweiz-und-international.html

2.	CoVariants. Accessed September 16, 2022. https://covariants.org/per-country?region=Switzerland
