## Supplementary figures and images for "Association of viral variant and vaccination status with the occurrence of symptoms compatible with post-acute sequelae after primary SARS-CoV-2 infection"

### Figure S1

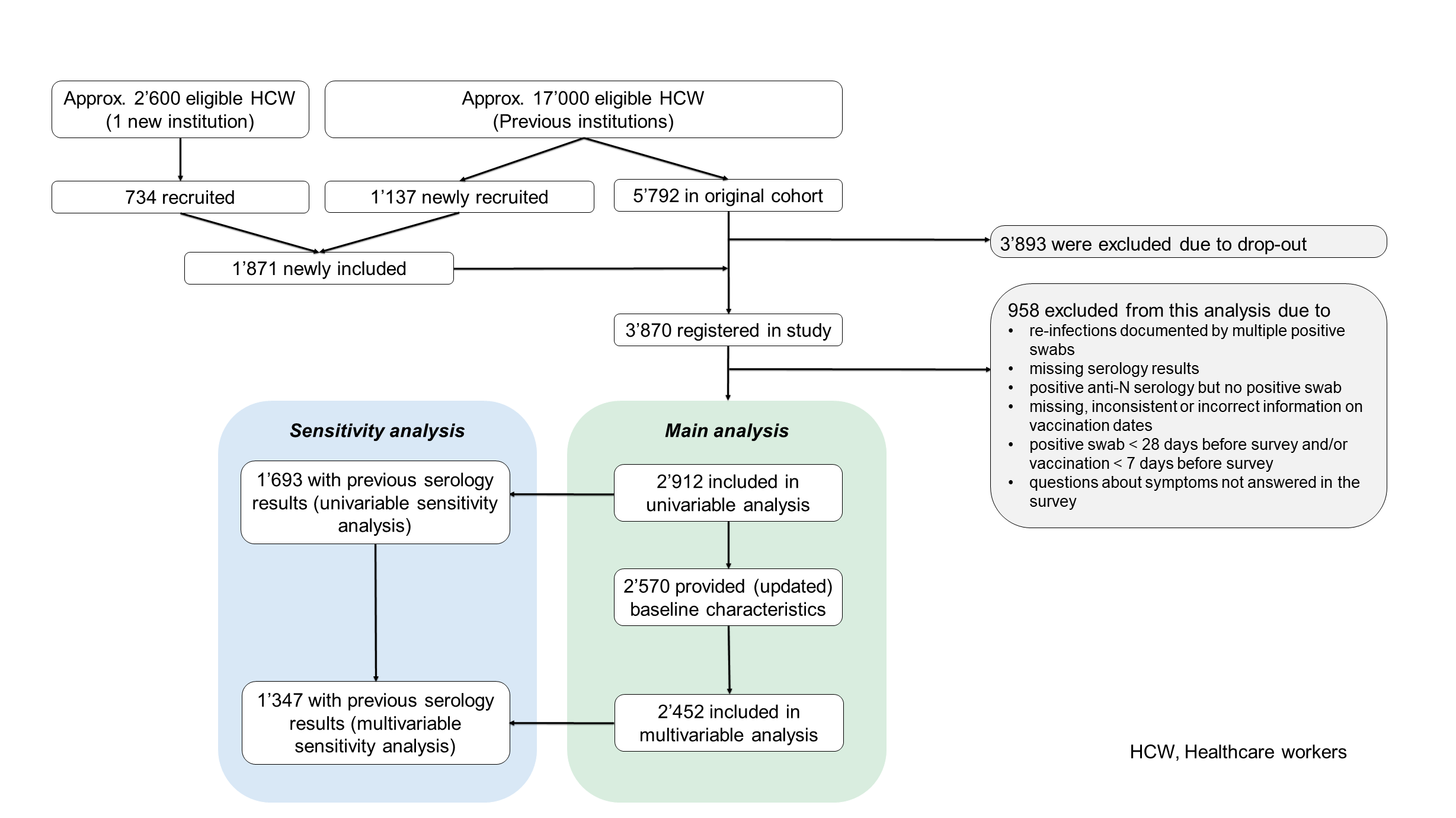

### Figure S2

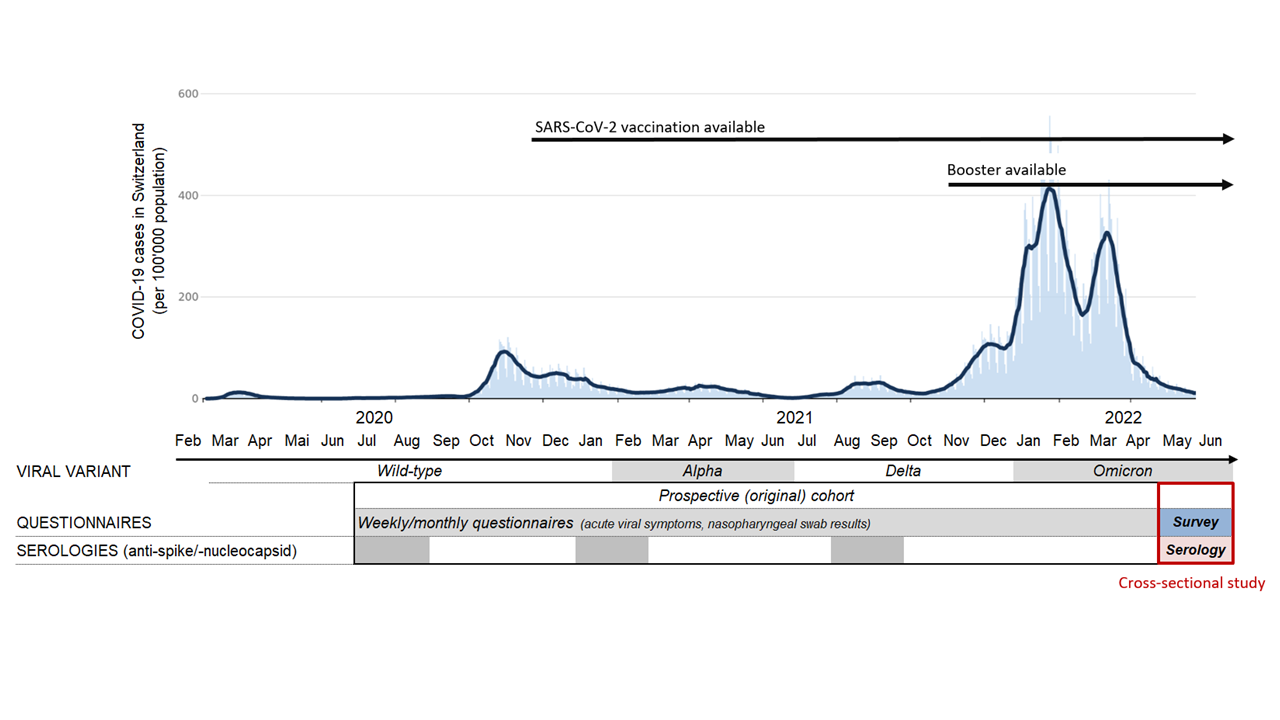

### Figure S3

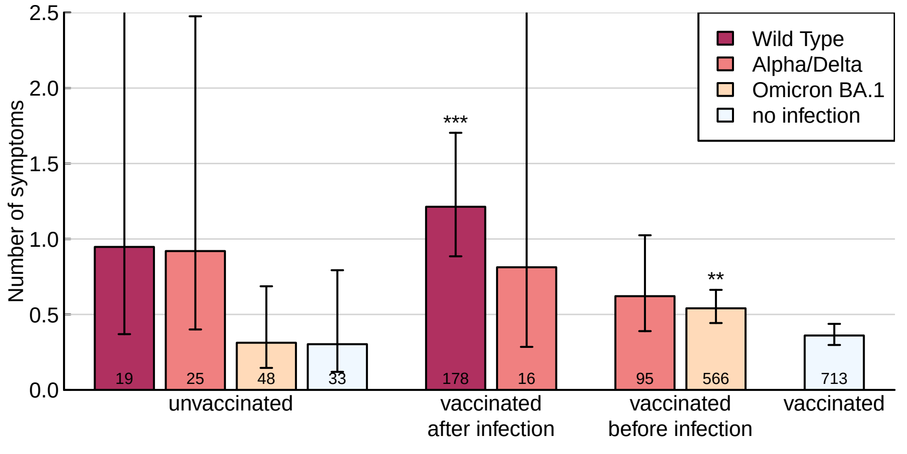

### Figure S4

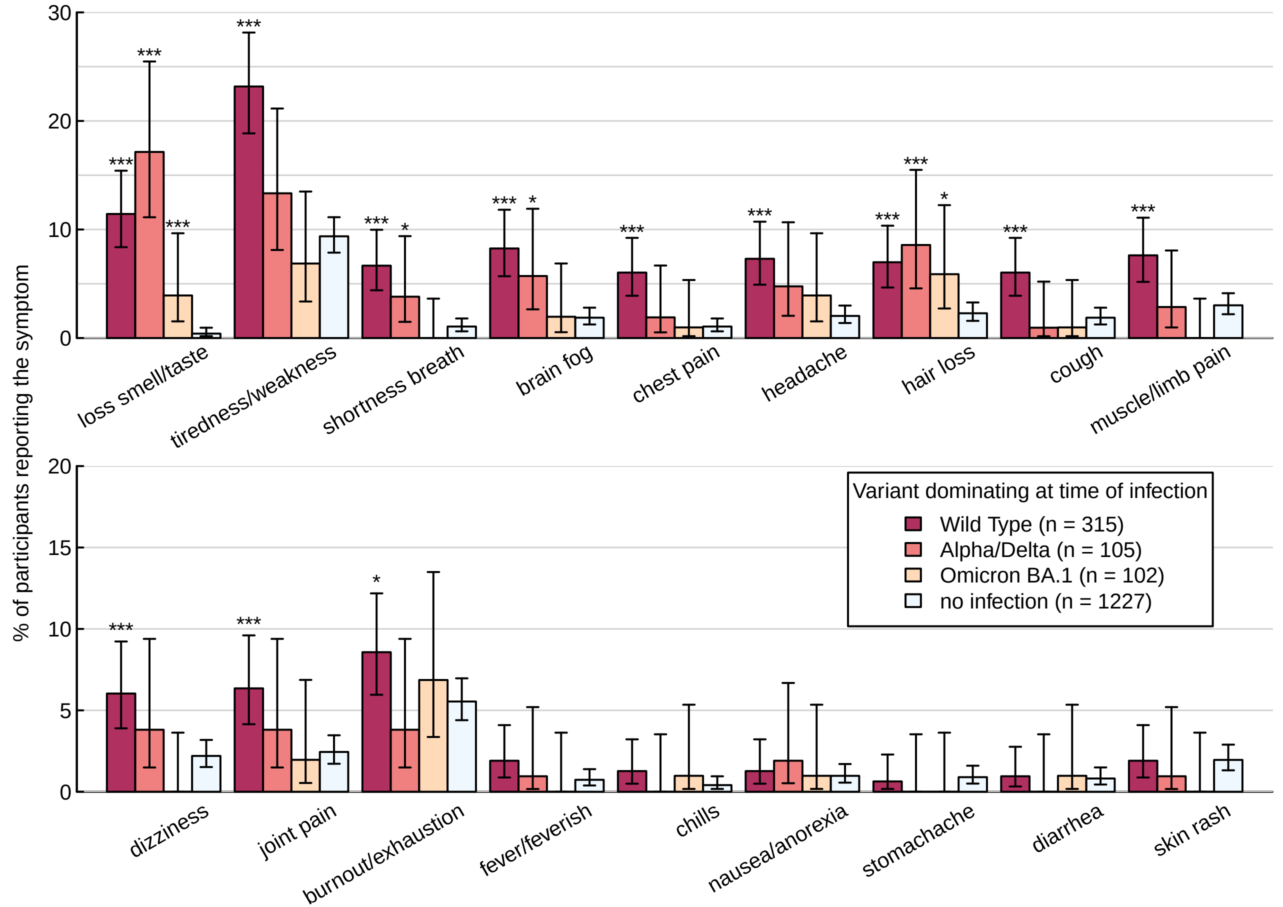

### Figure S5

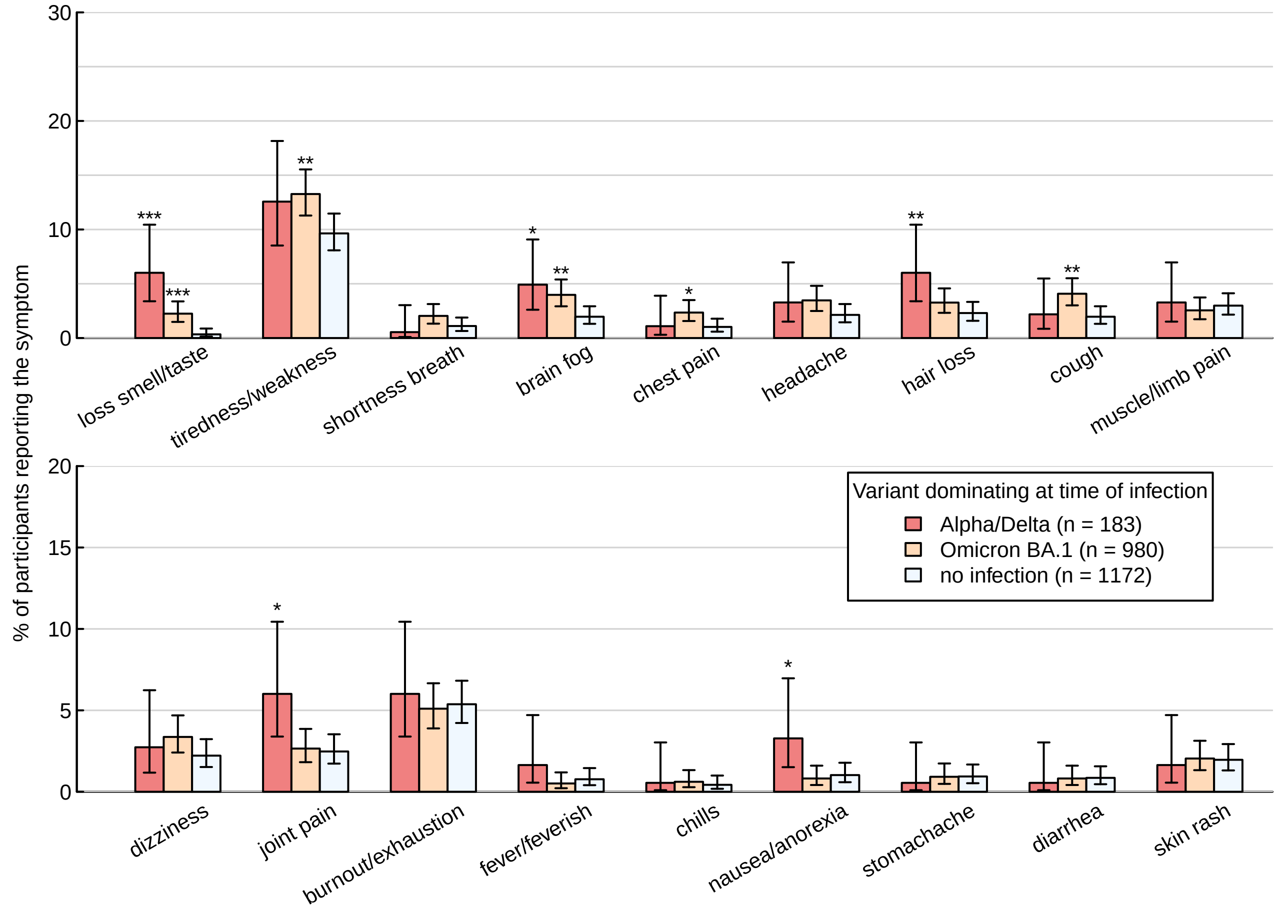

### Figure S6

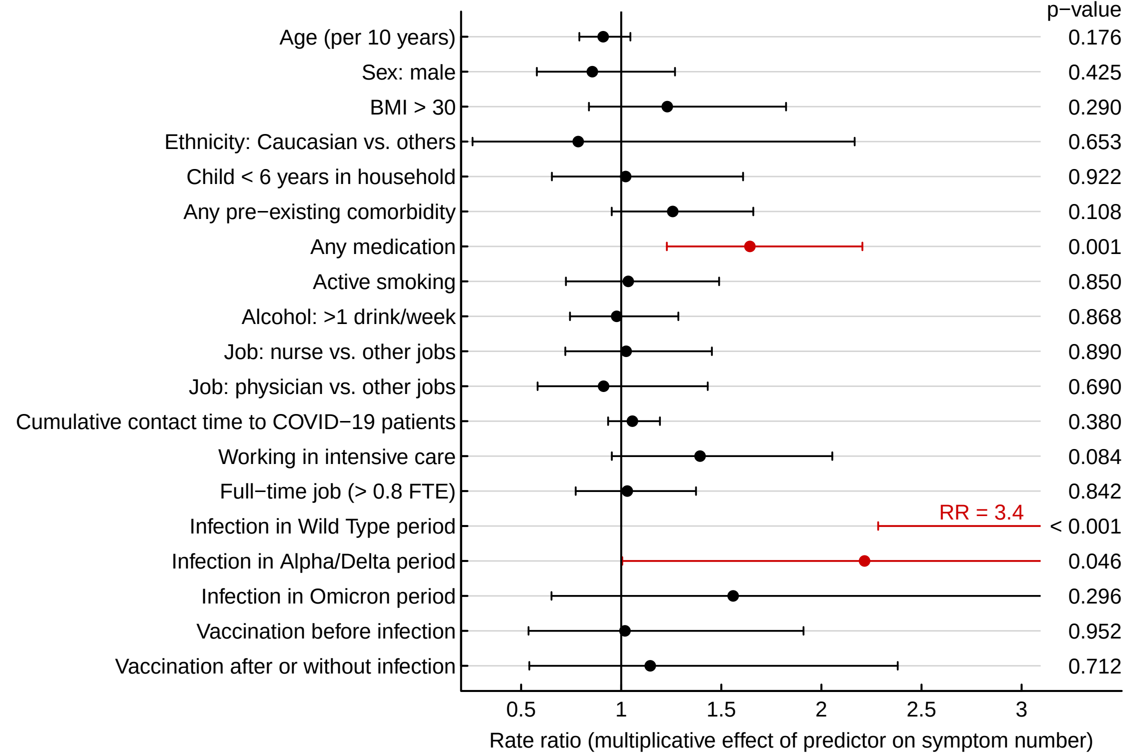
